# Factors associated with the uptake of intermittent preventive treatment in pregnancy with sulfadoxine–pyrimethamine: The experiences of postpartum women attending child welfare clinics in three rural districts in the Western Region of Ghana

**DOI:** 10.64898/2026.06.23.26355941

**Authors:** Gifty Amugi, Daniel Tetteh Agudey, Nana Mireku Gyimah, Pius Mensah, Isaac Kwarkye, Gabriel Yelkoang Gozie Yengliereh, Kofi Jean Claude, Bryony Brookman-Eshun, Sylvester Chinbuah

## Abstract

**Background:** Intermittent preventive treatment in pregnancy with sulfadoxine–pyrimethamine (IPTp-SP) is a key preventive strategy. However, optimal uptake remains inconsistent despite high antenatal care (ANC) attendance. This study assessed factors associated with IPTp-SP uptake and explored the experiences of postpartum women in rural Ghana.

**Methods:** A mixed-method study was conducted among 1,155 postpartum women attending child welfare clinics in Jomoro, Prestea-Huni Valley and Ellembelle districts of the Western Region of Ghana. Quantitative data were collected using structured questionnaires and analysed using descriptive statistics and chi-square tests. Qualitative data from in-depth interviews and focus group discussions were analysed thematically to explore women’s experiences and perceptions.

**Results:** Overall, 73.5% (812/1105) of respondents received at least three doses of SP during pregnancy, in line with WHO recommendations. The most common number of doses received was three doses (31.5%, 348/1105), followed by four doses (26.4%, 292/1105), while a smaller proportion (8.1%, 90/1105) received only one dose. Knowledge of malaria in pregnancy was generally high: 92.7% (1027/1155) of respondents correctly identified its mode of transmission, while 75.1% (830/1155) and 83.5% (923/1155) were aware of the effects of malaria on pregnancy and the foetus, respectively. Uptake was not significantly associated with socio-demographic characteristics, including age, education, occupation, marital status, gravidity, and parity (p > 0.05). However, number of ANC visits was significantly associated with uptake (p = 0.006). Although not statistically significant, lower uptake was observed among peri-urban residents and uninsured women. Qualitative findings indicated that while women recognized the benefits of IPTp-SP, side effects such as nausea, dizziness, and discomfort, as well as challenges with tablet formulation and dosing negatively influenced adherence.

**Conclusions:** IPTp-SP uptake was high and largely independent of socio-demographic factors but strongly influenced by ANC attendance. Addressing experiential barriers and strengthening patient-centered counselling during ANC may further improve uptake and adherence.

## Introduction

Malaria remains a major public health problem, particularly in sub–Saharan Africa (SSA). It is caused by protozoan parasites of the genus Plasmodium and transmitted through the bite of an infective female Anopheles mosquito. *Plasmodium falciparum* is responsible for most severe disease and deaths. Globally, an estimated 241 million malaria cases were reported in 2020 across 85 endemic countries, an increase from 227 million cases in 2019, with the majority occurring in the World Health Organization (WHO) African Region [1]. Each year, about 50 million women become pregnant in malaria endemic countries, and more than half reside in SSA where *P. falciparum* transmission is intense [2,3]. Malaria during pregnancy contributes substantially to maternal and neonatal morbidity and mortality. It is estimated that approximately 10,000 maternal deaths and 200,000 infant deaths occur annually due to malaria infection in pregnancy, largely driven by severe maternal anaemia, low birth weight, and preterm delivery [2,4]. Pregnant women and children under five years remain the most vulnerable groups in endemic settings [1,5]. Ghana is among the countries with stable malaria transmission throughout the year, with seasonal peaks in the northern zones. Malaria accounts for about 38 % of outpatient visits, 36 % of hospital admissions, and 33 % of deaths among children under five years. Among pregnant women, malaria contributes to about 14 % of outpatient cases, 11 % of hospital admissions, and 9 % of maternal deaths [6,7]. *Plasmodium falciparum* accounts for about 90 to 98 % of infections in Ghana, with *P. malariae* and *P. ovale* contributing a smaller proportion [7,8]. Historically, malaria prevention in pregnancy in SSA relied on weekly chloroquine prophylaxis during antenatal care. However, poor adherence and widespread parasite resistance limited its effectiveness. In 2000, the WHO recommended intermittent preventive treatment in pregnancy with sulfadoxine pyrimethamine (IPTp-SP) as a replacement strategy [9]. This strategy, delivered through routine antenatal care services, is one of the three pillars of malaria prevention in pregnancy, alongside insecticide treated nets and effective case management [1,9].

In 2012, the WHO updated its policy to recommend administration of sulfadoxine pyrimethamine (SP) at each scheduled antenatal care visit starting from the second trimester, with doses given at least one month apart until delivery [10]. Evidence shows that receiving three or more doses is associated with improved birth outcomes and reduced maternal anaemia [11,12].

Despite policy adoption, coverage of IPTp-SP remains sub optimal in many settings. In Ghana, the 2016 Demographic and Health Survey reported that 63 % of pregnant women received three or more doses, while 10 % received none [13]. In the Western Region, uptake of three or more doses has remained around 50 % in recent years, despite high antenatal attendance exceeding 70 % for four or more visits. The 2022 Demographic and Health Survey indicated that 49.1 % of women in the region received three or more doses during pregnancy [14]. This gap between antenatal attendance and optimal uptake suggests the presence of client level and health system factors that require further examination. Previous studies in Ghana and other parts of SSA have identified maternal age, education, parity, timing of antenatal initiation, knowledge of malaria in pregnancy, and health system factors as determinants of uptake [7,15–17]. However, limited evidence exists from rural districts in the Western Region that integrates quantitative assessment with lived experiences of postpartum women. Understanding both measurable predictors and experiential factors is essential to guide targeted interventions within routine maternal health services. This study therefore assessed factors associated with the uptake of IPTp-SP among postpartum women attending child welfare clinics in Jomoro, Prestea-Huni valley and Ellembelle districts of the Western Region of Ghana. Findings from this study will inform district and regional malaria control efforts aimed at improving protection of pregnant women and newborns in endemic settings.

## Methods

### Study design

The study was conducted using a mixed method approach. Quantitative and qualitative data were collected during the same period and analysed separately, then interpreted together. This study was conducted between August 7, 2024 and August 6, 2025.

### Study setting

The study was conducted in three rural districts in the Western Region of Ghana, namely Ellembelle, Prestea Huni Valley, and Jomoro. These districts are characterized by perennial malaria transmission with seasonal peaks linked to rainfall patterns. The region reports sustained malaria burden among pregnant women despite high antenatal care attendance. District health facilities include hospitals, health centres, and Community based Health Planning and Services compounds that provide routine antenatal and child welfare services.

### Study population

The study population comprised postpartum women who had delivered within three months prior to data collection and were attending child welfare clinics in selected facilities within the three districts.

#### Inclusion criteria

Women who delivered within three months before the study and attended antenatal care during their most recent pregnancy were eligible.

#### Exclusion criteria

Women who did not attend antenatal care during pregnancy and those with documented glucose-6-phosphate-dehydrogenase (G6PD) deficiency who were not eligible to receive sulfadoxine pyrimethamine were excluded.

### Sample size determination

For the quantitative component, sample size was estimated using Cochran’s formula for proportions with a 95% confidence level and 5% margin of error [18]. District specific coverage of three (3) or more doses of SP used as the estimated prevalence in Ellembelle, Prestea Huni Valley and Jomoro were 60.6%, 50.8% and 52.9% respectively [14]. The calculated sample sizes were therefore 367 for Ellembelle, 384 for Prestea Huni Valley, and 383 for Jomoro, giving a total sample size of 1134. To account for non-response, recruitment continued until 1155 eligible women were enrolled. For the qualitative component, a total of 28 in-depth interviews were conducted with postpartum women. Sample size was guided by the principle of data saturation as described in qualitative research literature [19,20].

### Sampling procedure

A multistage sampling approach was used for the quantitative survey. First, child welfare clinic sites were selected within each district using simple random sampling from the list of functional facilities obtained from district health directorates. Second, systematic sampling was applied on clinic days to recruit eligible women. After selecting the first participant at random, every third eligible woman was invited to participate until the required sample was achieved.

For the qualitative study, purposive sampling was employed to ensure representation of women who completed the recommended doses and those who did not. Separate focus group discussions were conducted for women who completed recommended doses and those who did not, to encourage open discussion and minimize social desirability bias.

### Data collection

#### Quantitative data

Data were collected using a structured interviewer administered questionnaire programmed in Open Data Kit (ODK). The tool captured socio demographic characteristics, antenatal attendance, number of doses received, timing of initiation, knowledge of malaria in pregnancy, and perceived barriers to uptake. The questionnaire was developed in English and administered in English or local languages as appropriate.

The tool was pretested in a district outside the study area using at least 10 percent of the estimated sample to assess clarity, flow, and internal consistency. Necessary revisions were made before final data collection.

#### Qualitative data

The qualitative component was designed to provide contextual understanding of the quantitative findings related to IPTp-SP uptake. Data were collected through focus group discussions and in-depth interviews using semi-structured interview guides developed from the study objectives and existing literature on malaria in pregnancy and IPTp-SP uptake. The guides explored women’s experiences with SP administration, perceptions of side effects, understanding of the benefits of IPTp-SP, provider communication, and barriers influencing adherence.

Three focus group discussions were conducted among postpartum women, with each group comprising eight participants. In addition, in-depth interviews were conducted with postpartum women, midwives, and pharmacists involved in antenatal care and SP administration. Interviews and discussions were conducted in Twi and Nzema by trained research assistants experienced in qualitative interviewing. All interviews were audio recorded with participants’ consent. Audio recordings were transcribed verbatim and translated into English. To ensure accuracy and preserve meaning, translated transcripts were independently reviewed and back translated where necessary. Data collection continued until thematic saturation was achieved, defined as the point at which no new themes or insights emerged from subsequent interviews.

### Study variables

The primary outcome variable was uptake of ITPp-SP, defined as receipt of at least one dose of SP during pregnancy.

Independent variables included age, educational level, occupation, marital status, residence, gravidity, parity, insurance status, gestational age at first dose, number of antenatal visits, knowledge of malaria in pregnancy, availability of SP, and perceived side effects.

### Data analysis

Quantitative data were exported to Statistical Package for Social Sciences version 21 for analysis. Descriptive statistics were computed as frequencies and proportions. Chi-square tests were used to assess associations between categorical independent variables and uptake of SP. Statistical significance was set at p less than 0.05.

Qualitative data were analysed using thematic analysis. Transcripts were read repeatedly to achieve familiarization with the data. Initial codes were generated manually and grouped into broader categories based on similarities and recurring patterns. Themes were then developed inductively from the coded data and refined through iterative discussions among the research team. Credibility of findings was enhanced through investigator review, comparison of responses across participant groups, and triangulation with quantitative findings. Findings were integrated with quantitative results during interpretation.

### Ethical considerations

Ethical approval was obtained from the Ghana Health Service Ethical Review Committee (GHS-ERC:021/03/24). Permission was also obtained from district health directorates and facility managers. Written informed consent was obtained from all participants. Confidentiality was maintained by using unique identifiers and storing data securely. Participation was voluntary and respondents were free to withdraw at any stage without consequence.

## Results

### Socio-demographic and obstetric characteristics of post-partum women included in the study

The study included 1155 post-partum women, most of whom were aged 25–29 years (29.8%). Over half of the participants were married (55.5%) and the majority resided in rural areas (67.6%). In terms of education, secondary education was the most common level attained (37.2%), while 18.3% had no formal education. Most women (93.7%) were enrolled in the national health insurance scheme, 74.5% initiated SP at 16 weeks of gestation and 42.9% attended four to six antenatal care visits during pregnancy (Table 1).

**Table 1:**
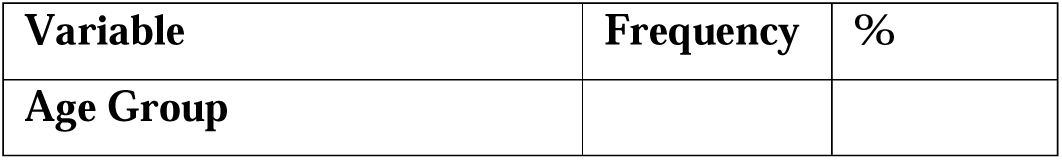
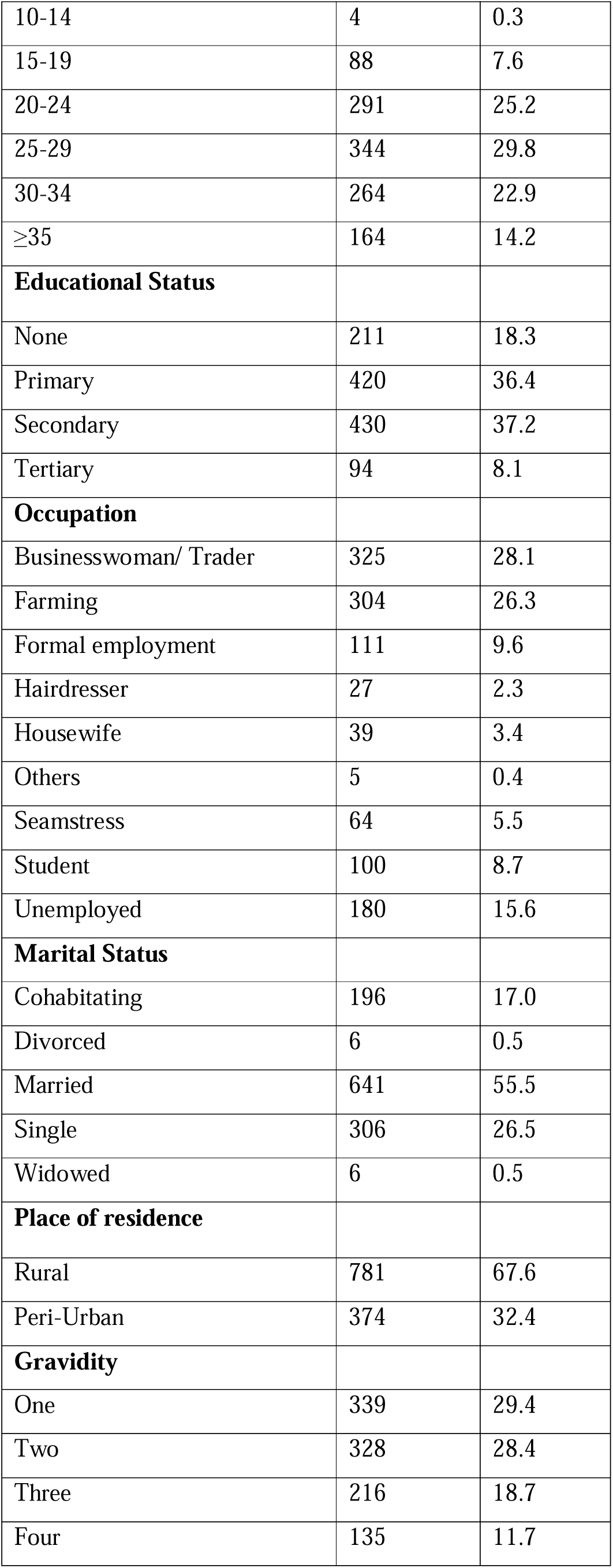
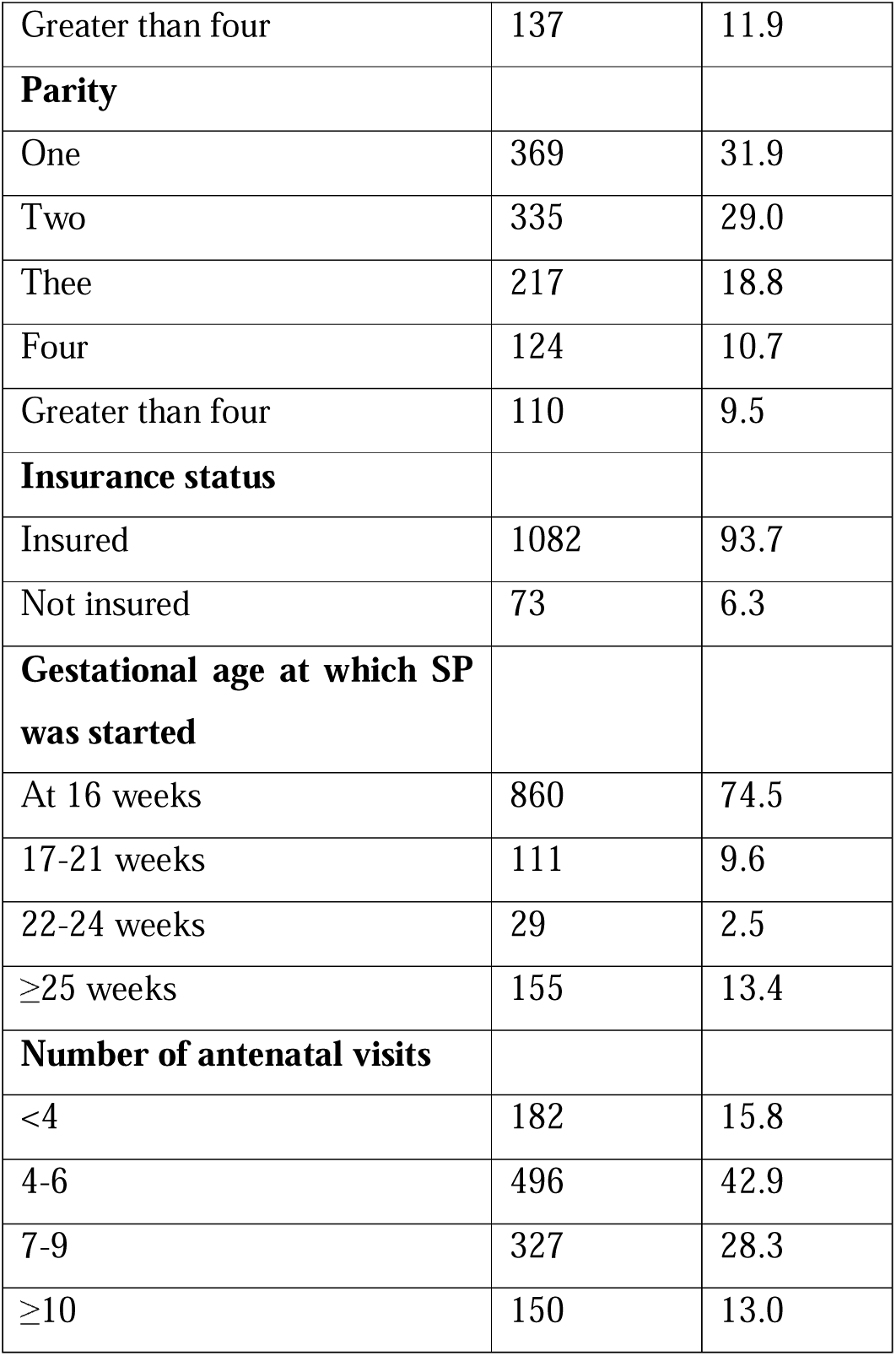
Socio-demographic and obstetric characteristics of post-partum women attending Child Welfare Clinics (CWC) in three rural districts in the Western Region of Ghana.

| Variable | Frequency | % |
| --- | --- | --- |
| Age Group |  |  |
| 10-14 | 4 | 0.3 |
| 15-19 | 88 | 7.6 |
| 20-24 | 291 | 25.2 |
| 25-29 | 344 | 29.8 |
| 30-34 | 264 | 22.9 |
| ≥35 | 164 | 14.2 |
| **Educational Status** |  |  |
| None | 211 | 18.3 |
| Primary | 420 | 36.4 |
| Secondary | 430 | 37.2 |
| Tertiary | 94 | 8.1 |
| **Occupation** |  |  |
| Businesswoman/ Trader | 325 | 28.1 |
| Farming | 304 | 26.3 |
| Formal employment | 111 | 9.6 |
| Hairdresser | 27 | 2.3 |
| Housewife | 39 | 3.4 |
| Others | 5 | 0.4 |
| Seamstress | 64 | 5.5 |
| Student | 100 | 8.7 |
| Unemployed | 180 | 15.6 |
| **Marital Status** |  |  |
| Cohabiting | 196 | 17.0 |
| Divorced | 6 | 0.5 |
| Married | 641 | 55.5 |
| Single | 306 | 26.5 |
| Widowed | 6 | 0.5 |
| **Place of residence** |  |  |
| Rural | 781 | 67.6 |
| Peri-Urban | 374 | 32.4 |
| **Gravidity** |  |  |
| One | 339 | 29.4 |
| Two | 328 | 28.4 |
| Three | 216 | 18.7 |
| Four | 135 | 11.7 |
| Greater than four | 137 | 11.9 |
| **Parity** |  |  |
| One | 369 | 31.9 |
| Two | 335 | 29.0 |
| Three | 217 | 18.8 |
| Four | 124 | 10.7 |
| Greater than four | 110 | 9.5 |
| **Insurance status** |  |  |
| Insured | 1082 | 93.7 |
| Not insured | 73 | 6.3 |
| **Gestational age at which SP was started** |  |  |
| At 16 weeks | 860 | 74.5 |
| 17-21 weeks | 111 | 9.6 |
| 22-24 weeks | 29 | 2.5 |
| ≥25 weeks | 155 | 13.4 |
| **Number of antenatal visits** |  |  |
| <4 | 182 | 15.8 |
| 4-6 | 496 | 42.9 |
| 7-9 | 327 | 28.3 |
| ≥10 | 150 | 13.0 |

### The proportion of post-partum women who took IPTp during pregnancy

About 95.7% (1105/1155) of respondents reported taking IPTp-SP during pregnancy (Fig 2).

**Fig 1:**
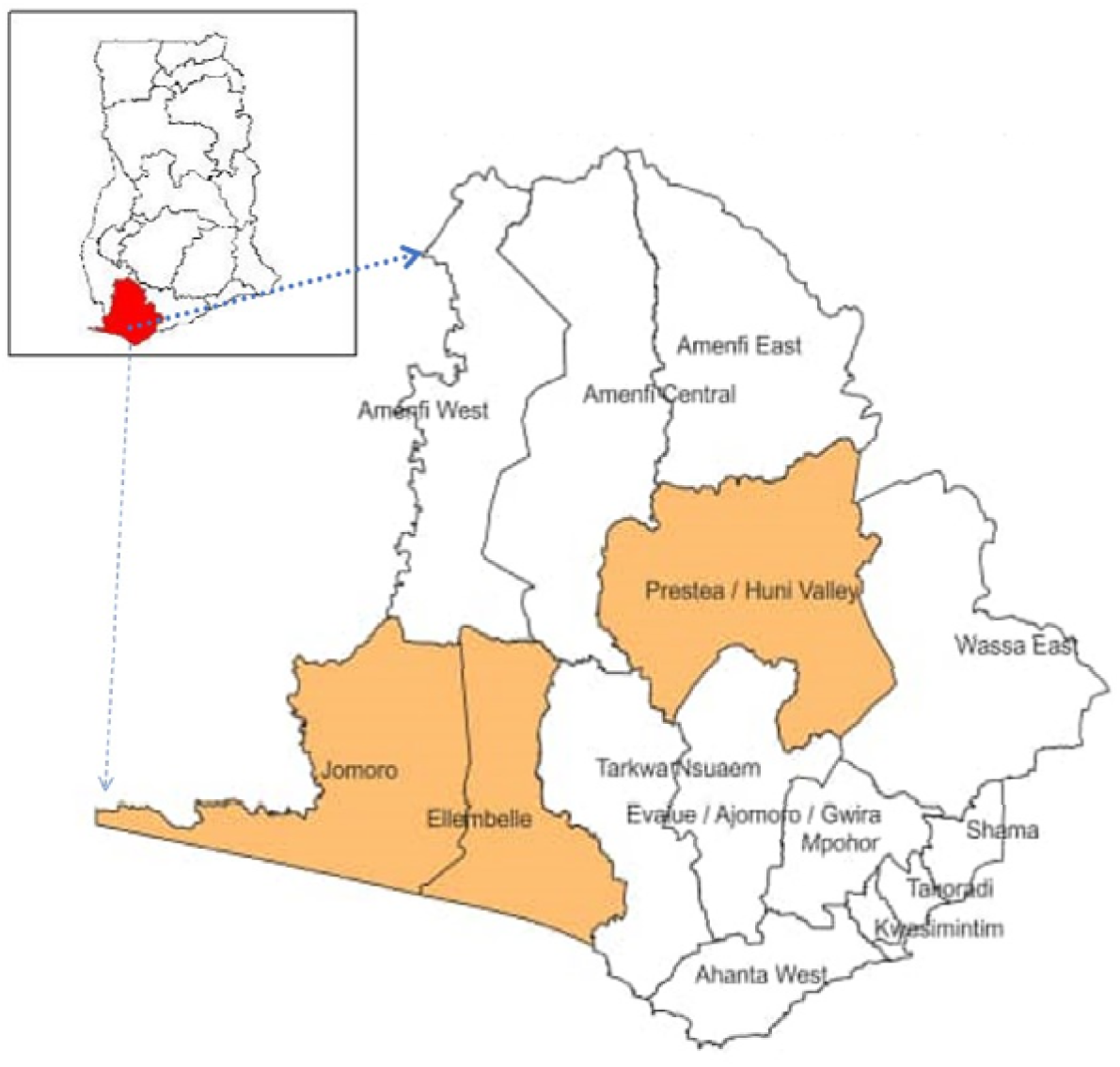
Map of Western Region showing the three study districts

**Fig 2:**
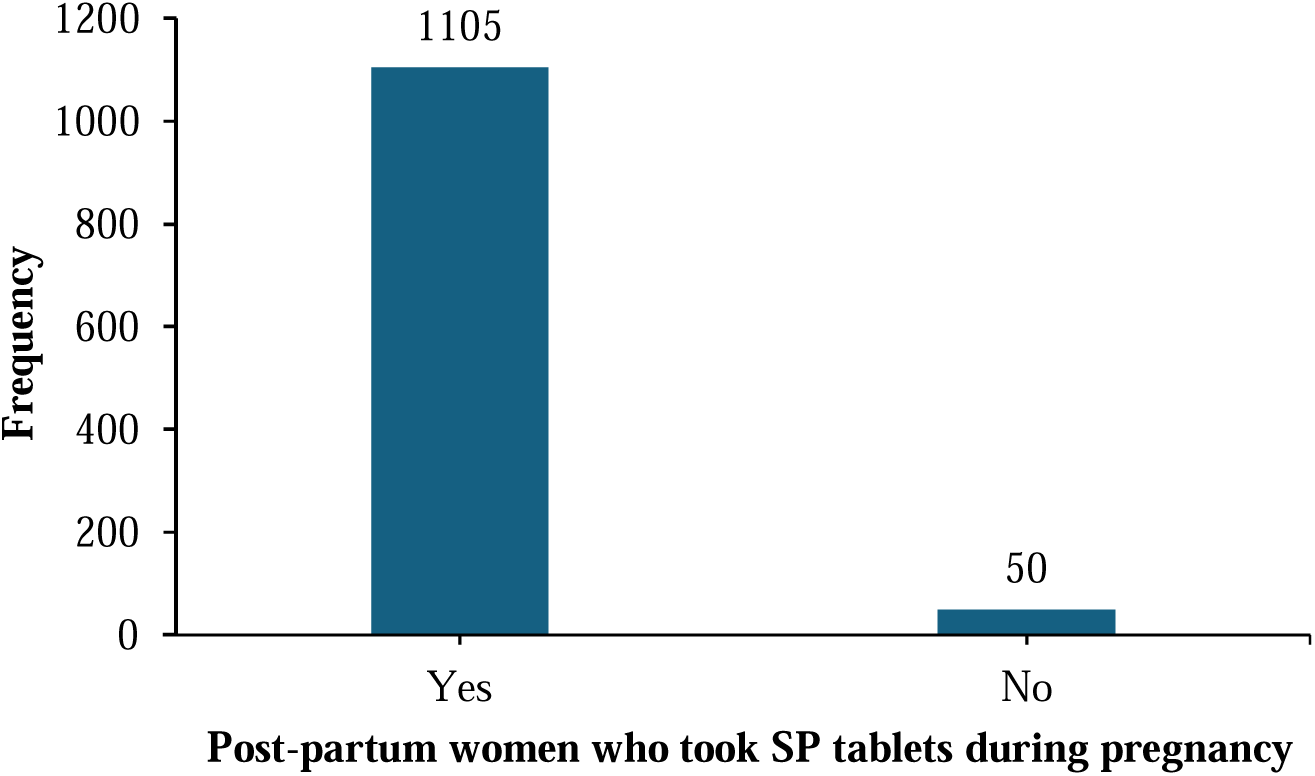
Post-partum women who took IPTp during pregnancy

### The level of knowledge of Postpartum Women on Malaria in Pregnancy (MIP) And IPTP-SP

Table 2 shows the knowledge of respondents on malaria in pregnancy and IPTp-SP. It was found that 92.7% (1027/1155) of respondents have knowledge on how malaria is transmitted in pregnancy, 75.1% (830/1155) had knowledge of the effect of malaria on pregnancy, 83.5% (923/1155) had knowledge on the effect of malaria infection on the foetus and 91.7% (1013/1155) had knowledge of prevention of malaria infection in pregnancy.

**Table 2:** Knowledge Of Postpartum Women on MIP And IPTP-SP.

| Knowledge of MIP | Yes (n) | % | No (n) | % |
| --- | --- | --- | --- | --- |
| Knowledge of how malaria is transmitted in pregnancy. | 1027 | 92.9% | 78 | 7.1% |
| Knowledge of the effect of malaria on pregnancy. | 830 | 75.1% | 275 | 24.9% |
| Knowledge of the effect of malaria infection on the fetus. | 923 | 83.5% | 182 | 16.5% |
| Knowledge of prevention of malaria infection in pregnancy. | 1013 | 91.7% | 92 | 8.3% |

### Causes of Malaria

About 89.2% (986/1105) of participants identified mosquito bites as the main cause, while only a small number attributed malaria to incorrect causes such as exposure to heat or drinking dirty water, and some reported having no idea (Fig 3).

**Fig 3:**
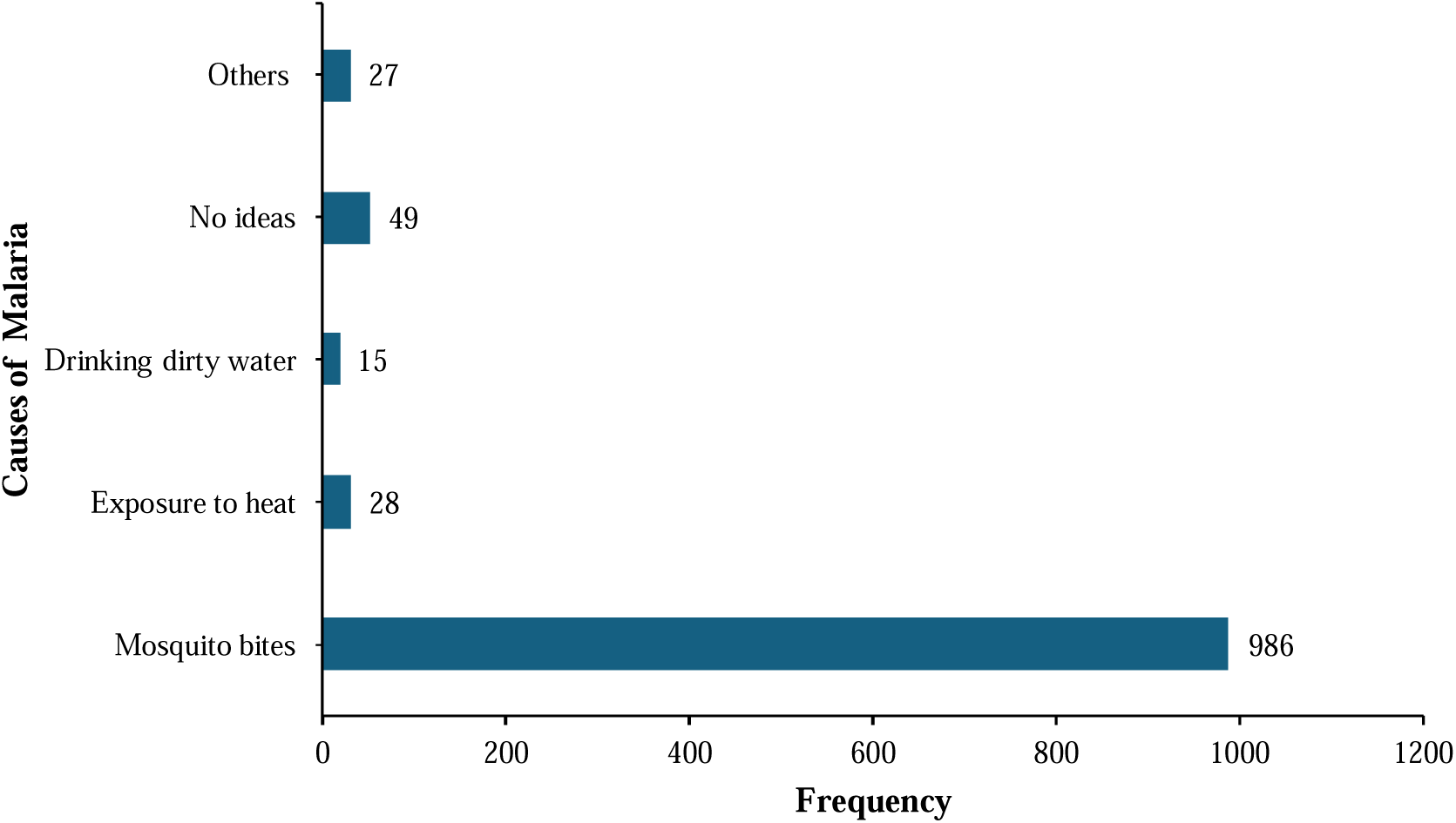
Level of knowledge on the Causes Of Malaria among Postpartum Women

### Factors affecting the uptake of SP

Side effects associated with SP were the most frequently reported factor influencing the uptake of IPTp SP among respondents (64%, 32/50) (Fig 4).

**Fig 4:**
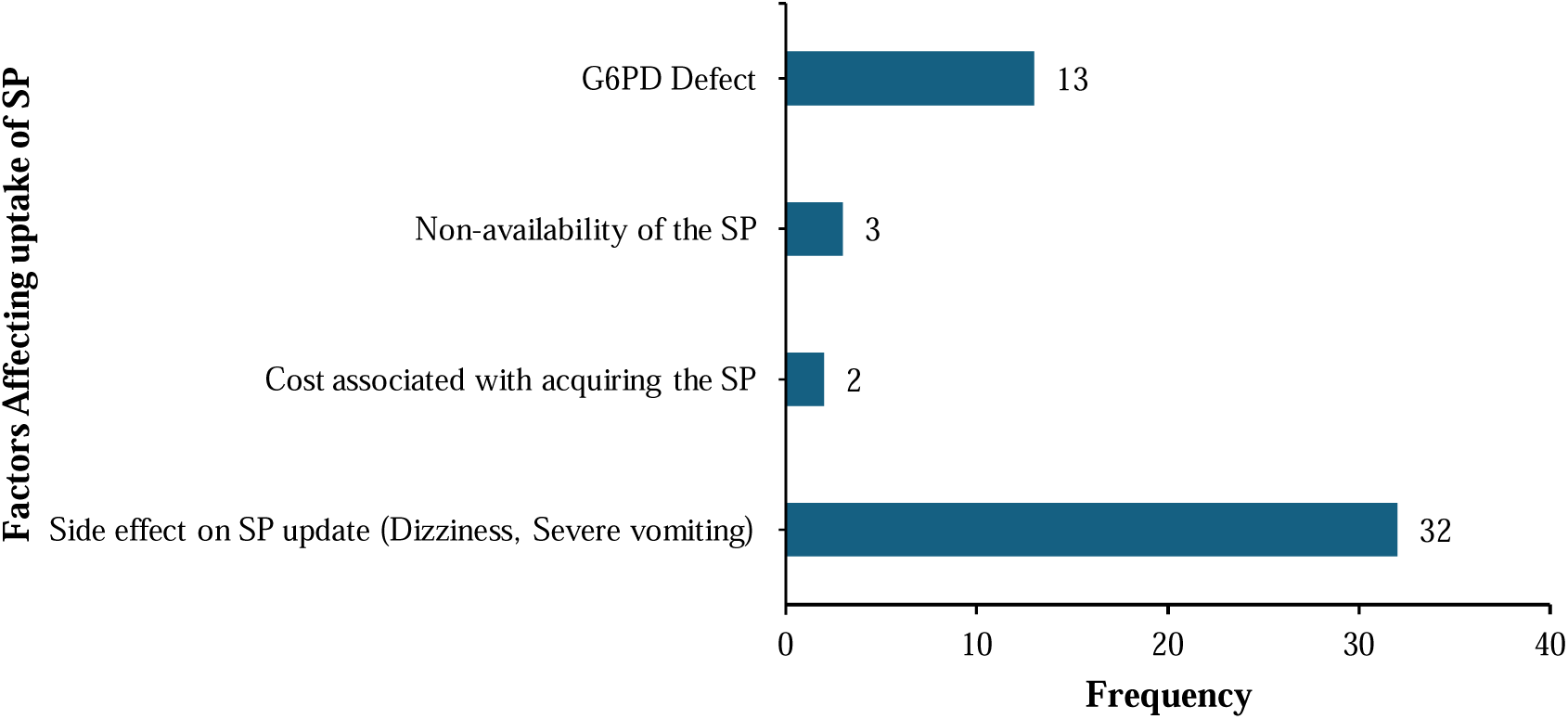
Factors influencing uptake of SP among postpartum women who did not take SP

### Number of IPTp SP doses received during pregnancy among postpartum women

About 31.5% (348/1105) of respondents received three doses of SP during pregnancy followed by four doses (26.4, 292/1105). However, 8.1% (90/1105) of women received only one dose (Fig 5). Those who received at least three doses as recommended by WHO were about 73.5% (812/1105).

**Fig 5:**
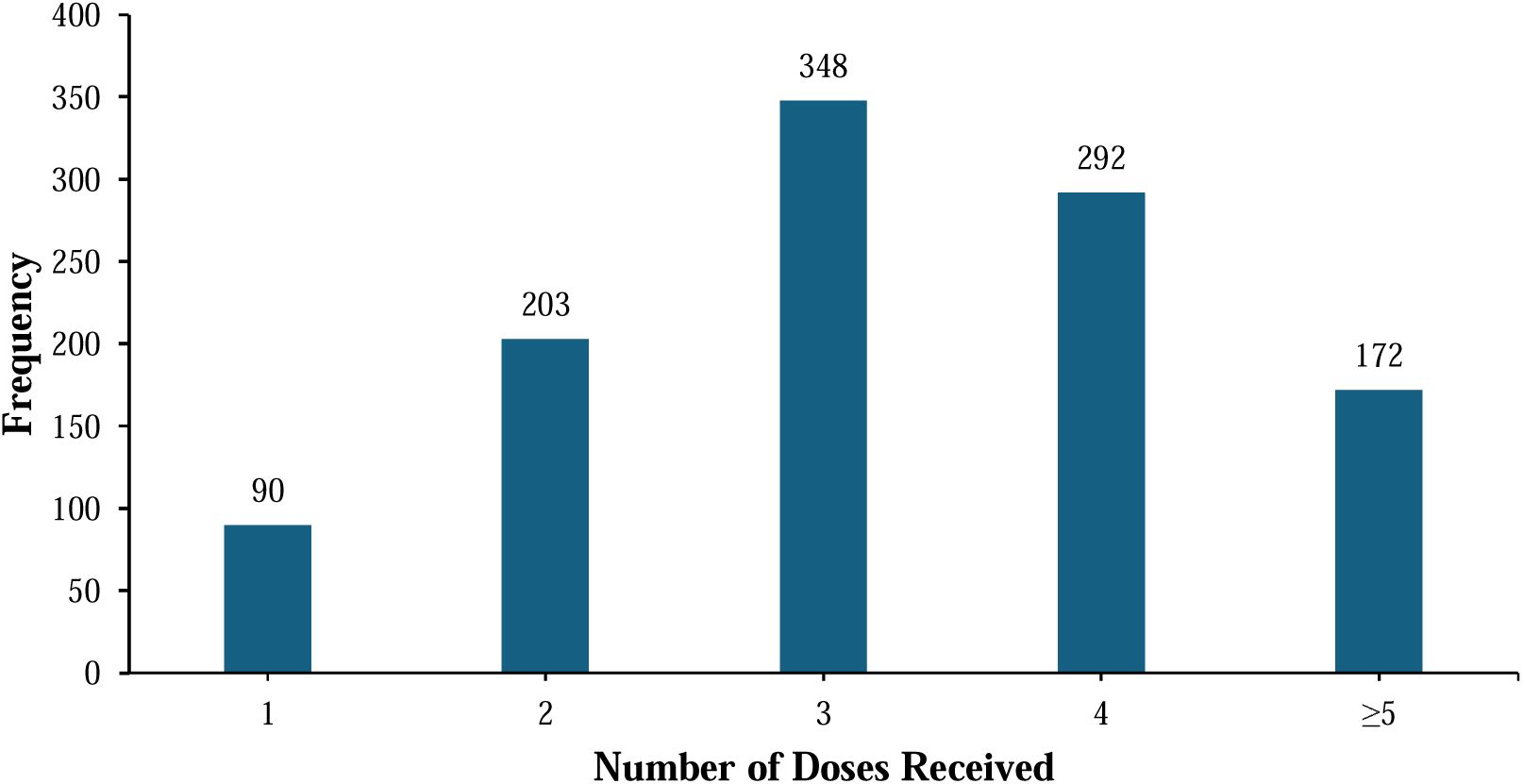
Distribution of the number of IPTp SP doses received during pregnancy among postpartum women

### Association Between Uptake of IPTp-SP During Pregnancy and Selected Socio-Demographic and Obstetric Characteristics Among Postpartum Women

The number of antenatal care (ANC) visits was significantly associated with IPTp-SP uptake (p = 0.006). Women who attended fewer than four ANC visits had a comparatively lower uptake (91%) and higher non-use (9%), whereas those with 4–6 visits demonstrated the highest uptake (97%). There was no statistically significant association between IPTp-SP uptake and key demographic factors such as age (p = 0.153), educational status (p = 0.882), occupation (p = 0.907), marital status (p = 0.228), gravidity (p = 0.458), or parity (p = 0.601) (Table 3).

**Table 3:** Association Between Uptake of IPTp-SP During Pregnancy and Selected Socio-Demographic and Obstetric Characteristics Among Postpartum Women.

| Variable | Proportion of post-partum women who took IPTp-SP during pregnancy |  |  |  |  | p-value |
| --- | --- | --- | --- | --- | --- | --- |
|  | Yes | % | No | % | Total |  |
| **Age Group** |  |  |  |  |  | 0.153 |
| 10-14 | 2 | 50% | 2 | 50% | 4 |  |
| 15-19 | 85 | 97% | 3 | 3% | 88 |  |
| 20-24 | 279 | 96% | 12 | 4% | 291 |  |
| 25-29 | 330 | 96% | 14 | 4% | 344 |  |
| 30-34 | 254 | 96% | 10 | 4% | 264 |  |
| ≥35 | 155 | 95% | 9 | 5% | 164 |  |
| **Educational Status** |  |  |  |  |  | 0.882 |
| None | 201 | 95% | 10 | 5% | 211 |  |
| Primary | 400 | 95% | 20 | 5% | 420 |  |
| Secondary | 414 | 96% | 16 | 4% | 430 |  |
| Tertiary | 90 | 96% | 4 | 4% | 94 |  |
| **Occupation** |  |  |  |  |  | 0.907 |
| Businesswoman/<br>Trader | 311 | 96% | 14 | 4% | 325 |  |
| Farming | 290 | 95% | 14 | 5% | 304 |  |
| Formal<br>employment | 108 | 97% | 3 | 3% | 111 |  |
| Hairdresser | 27 | 100% | 0 | 0% | 27 |  |
| Housewife | 38 | 97% | 1 | 3% | 39 |  |
| Others | 5 | 100% | 0 | 0% | 5 |  |
| Seamstress | 61 | 95% | 3 | 5% | 64 |  |
| Student | 94 | 94% | 6 | 6% | 100 |  |
| Unemployed | 171 | 95% | 9 | 5% | 180 |  |
| **Marital Status** |  |  |  |  |  | 0.228 |
| Cohabiting | 183 | 93% | 13 | 7% | 196 |  |
| Divorced | 5 | 83% | 1 | 17% | 6 |  |
| Married | 617 | 96% | 24 | 4% | 641 |  |
| Single | 294 | 96% | 12 | 4% | 306 |  |
| Widowed | 6 | 100% | 0 | 0% | 6 |  |
| **Place of residence** |  |  |  |  |  | 0.073 |
| Rural | 753 | 96% | 28 | 4% | 781 |  |
| Peri-urban | 352 | 94% | 22 | 6% | 374 |  |
| **Gravidity** |  |  |  |  |  | 0.458 |
| One | 322 | 95% | 17 | 5% | 339 |  |
| Two | 318 | 97% | 10 | 3% | 328 |  |
| Three | 207 | 96% | 9 | 4% | 216 |  |
| Four | 126 | 93% | 9 | 7% | 135 |  |
| Greater than four | 132 | 96% | 5 | 4% | 137 |  |
| **Parity** |  |  |  |  |  | 0.601 |
| One | 350 | 95% | 19 | 5% | 369 |  |
| Two | 325 | 97% | 10 | 3% | 335 |  |
| Three | 207 | 95% | 10 | 5% | 217 |  |
| Four | 117 | 94% | 7 | 6% | 124 |  |
| Greater than four | 106 | 96% | 4 | 4% | 110 |  |
| **Insurance status** |  |  |  |  |  | 0.092 |
| Insured | 1038 | 96% | 44 | 4% | 1082 |  |
| Not insured | 67 | 92% | 6 | 8% | 73 |  |
| **Gestational age at**<br>**which SP was**<br>**received** |  |  |  |  |  | 0.152 |
| At 16 weeks | 816 | 95% | 44 | 5% | 860 |  |
| 17-21 weeks | 109 | 98% | 2 | 2% | 111 |  |
| 22-24 weeks | 29 | 100% | 0 | 0% | 29 |  |
| ≥25 weeks | 151 | 97% | 4 | 3% | 155 |  |
| **Number of**<br>**antenatal visits** |  |  |  |  |  | 0.006 |
| <4 | 166 | 91% | 16 | 9% | 182 |  |
| 4-6 | 483 | 97% | 13 | 3% | 496 |  |
| 7-9 | 313 | 96% | 14 | 4% | 327 |  |
| ≥10 | 143 | 95% | 7 | 5% | 150 |  |

### The experiences of post-partum women with IPTp-SP

#### Participants’ description

These qualitative findings describe the experiences and perceptions of postpartum women regarding IPTp-SP in Jomoro, Prestea-Huni Valley and Ellembelle districts of the Western Region of Ghana. Twenty-eight (28) postpartum women took part in focus group discussions and in-depth interviews. Nineteen (19) participants were aged 25 to 39 years and nine (9) were adolescents between the ages of 17 to 20 years.

Four main themes emerged from the analysis: discomfort after taking SP, awareness of the benefits of SP, knowledge of the harmful effects of malaria on the unborn child, and concerns about the number and formulation of SP tablets.

#### Experienced discomfort when SP was taken

Many participants reported unpleasant physical reactions after taking SP during antenatal care visits. Symptoms such as dizziness, weakness, nausea, and abdominal discomfort were commonly mentioned and sometimes created reluctance to attend clinic visits.

One participant explained, “I felt weak, dizzy and had the urge to vomit anytime I took the SP tablets during my ANC visits” (SS, 25-29 years). Another added, “I had continuous abdominal pains and nausea some few hours after taking the medication and this made me very uncomfortable throughout the day” (MA, 30-34 years).

Participants also expressed dissatisfaction with the requirement to chew the tablets during directly observed therapy. As one woman noted, “I found it very difficult chewing the 3 tablets. I think it is easy swallowing medicines, but the midwife always insisted I chew them” (AS, 35-39 years). In some cases, the anticipated discomfort discouraged clinic attendance. A younger participant stated, “There were times I did not want to attend ANC when my time was due to visit because of the thought of discomfort expected after taking the medicine” (DW, 15-19 years).

### Knowledge of the benefits of SP

Despite the discomfort reported, most women demonstrated a clear understanding of the preventive role of SP during pregnancy. Information provided during antenatal care visits was cited as the main source of this knowledge.

One respondent explained, “I was informed that the medicine served as a barrier and would protect my unborn baby from harm by the malaria parasites” (BD, 25-29 years). Another participant noted, “I know that the medicine protects pregnant women from malaria infection and made me healthy throughout my pregnancy” (NM, 35-39 years). Similarly, a participant emphasized the importance of preventing anaemia during pregnancy, stating, “I did not want to be anaemic; I learnt malaria in pregnancy can reduce the iron stores in pregnant women” (BE, 35-39 years).

### Awareness of the negative impact of malaria on unborn babies

Participants also demonstrated awareness of the potential adverse effects of malaria during pregnancy on the unborn child. Many linked malaria infections with complications affecting fetal growth and development.

A participant stated, “I understood that my unborn baby could be affected by the malaria parasites causing complications such as brain disorders during its development” (FC, 25-29 years). Another explained, “I learned that when my unborn baby is infected by the malaria parasites, it would lead to low birth weight which I did not want. I like big and strong babies” (MB, 35-39 years).

#### Concerns about the number and formulation of SP tablets

Several participants raised concerns about the number of tablets required per dose and the repeated dosing schedule during pregnancy. Suggestions were made for modifications that could make the medication easier to take.

One adolescent participant proposed, “If possible, the 3 tablets should be put together as one tablet so that they are taken once at a go during a visit” (TA, 15-19 years). Another suggested a different formulation, stating, “I think it will be very helpful if the tablets are changed into capsules so that they can be swallowed instead of chewing to make it easy for pregnant women to take” (AE, 20-24 years).

Some participants also recommended reducing the number of doses administered during pregnancy. As one woman noted, “Can the number of doses be reduced from 5 to 3 throughout the 9 months of pregnancy?” (SQ, 40-44 years), while another suggested a longer interval between doses: “Authorities should consider a dose every two months than a dose each month till delivery” (CK, 35-39 years).

## Discussion

This study examined the uptake of IPTp-SP among postpartum women in three rural districts in the Western Region of Ghana and explored the contextual factors influencing utilization through women’s lived experiences.

The study found that 96% of postpartum women received at least one dose of IPTp-SP during pregnancy, suggesting generally high coverage and acceptance of the intervention within the study districts. This finding is comparable to the 98.5% coverage reported by Ibrahim et al. (2017) in the Sunyani Municipality of Ghana [15]. The quantitative findings further showed that most women attended four or more ANC visits, which likely contributed to the high uptake observed. However, qualitative findings revealed that high uptake did not necessarily translate into positive experiences with SP administration. Many women described adverse reactions such as dizziness, nausea, abdominal discomfort, weakness and vomiting after taking SP. These experiences help explain why a minority of women failed to initiate or complete IPTp-SP despite attending ANC services. Similar findings were reported by Dun-Dery et al. (2021), who observed that unpleasant side effects and previous negative experiences reduced willingness to continue SP use during pregnancy [17].

This therefore suggests that while programmatic access to IPTp-SP may be high, experiential barriers continue to influence adherence behaviours. Some participants reported intentionally delaying or missing ANC visits due to fear of discomfort after SP administration. This finding is important because it demonstrates that utilization cannot be understood solely from coverage indicators without considering women’s perceptions and experiences during service delivery.

The study also demonstrated a statistically significant association between the number of ANC visits and IPTp-SP uptake (p = 0.006). Women who attended ANC more frequently were more likely to receive at least the recommended three doses of SP. Approximately 74% of respondents received three or more doses, exceeding the 42.4% reported by Agyeman et al. (2023) in Northern Ghana [21]. Similar associations between ANC attendance and IPTp-SP uptake have been documented in other Ghanaian studies [15,16]. Qualitative findings reinforced this association by showing that repeated ANC interactions improved women’s understanding of malaria prevention, increased trust in healthcare providers, and encouraged continued uptake of SP despite unpleasant side effects. Several women indicated that explanations given by midwives during ANC visits motivated them to continue taking SP because they understood its protective benefits for both mother and baby.

Interestingly, gestational age at initiation of SP was not significantly associated with uptake in this study. This contrasts with findings by Kumah et al. (2022), who reported that earlier initiation predicted higher completion rates [16]. The interviews provide additional context for this observation. Women who attended ANC consistently appeared more likely to continue receiving SP irrespective of when they initiated treatment, suggesting that continuity of ANC engagement may be more important than timing of initiation alone within this setting. Sociodemographic factors including age, education, occupation, marital status, parity, and residence were not significantly associated with IPTp-SP uptake. These findings are consistent with Ibrahim et al. (2017) but differ from studies that identified educational status as an important predictor of uptake [15,16]. The qualitative findings offer a possible explanation for this lack of association. Across educational and occupational groups, many women reported relying heavily on provider instruction and trust in health workers rather than personal knowledge in deciding to take SP. This suggests that within these rural settings, provider influence and routine ANC practices may have a stronger effect on uptake than individual socioeconomic characteristics.

Although general awareness of malaria transmission and prevention was high in the quantitative analysis, interviews revealed important gaps in detailed understanding of IPTp-SP. Many participants knew malaria could harm pregnancy and unborn babies, yet several could not identify SP by name or clearly explain its preventive role. Some women reported taking the medication primarily because it was administered during ANC visits rather than through informed understanding of its purpose. Similar observations have been reported by Nwaefuna et al. (2023) and De-Gaulle et al. (2022), who found that awareness of malaria in pregnancy did not always correspond with adequate knowledge of IPTp-SP [11,12].

The findings from this study therefore highlights an important distinction between compliance and informed adherence. While women generally complied with SP administration, the qualitative findings suggest that adherence was often dependent on provider direction and continued supervision. This may present challenges for long-term sustainability if provider communication is inadequate or if women experience adverse reactions without proper counselling and reassurance.

The study additionally identified concerns regarding the formulation and administration of SP. Some participants disliked chewing the tablets during directly observed therapy and preferred swallowing or alternative formulations such as capsules. Others expressed concern about the number of tablets and repeated monthly dosing schedule. Similar concerns regarding bitterness, side effects, and tablet burden have been documented in studies from Uganda and other malaria endemic settings [22]. These findings emphasize the need for more patient-centered approaches within ANC services, including improved counselling on expected side effects, supportive communication during directly observed therapy, and reassurance regarding the benefits of IPTp-SP.

Addressing these experiential and behavioural factors alongside routine service delivery may further strengthen IPTp-SP utilization and improve maternal and neonatal health outcomes in malaria endemic settings.

### Limitations and Strengths of the Study

The study’s retrospective design introduces potential recall bias, as responses were based on participants’ recollections of ANC experiences. Additionally, while place of residence was captured, the study did not explore the burden and cost of travel associated with ANC attendance, which could have influenced IPTp-SP uptake among rural women. Nonetheless, the study’s strength lies in its integration of both quantitative and experiential perspectives, providing a comprehensive understanding of IPTp-SP uptake dynamics in a real-world rural context.

## Conclusion

This study demonstrates that uptake of IPTp-SP among postpartum women in rural districts of the Western Region of Ghana is high, with most women receiving the recommended dose during pregnancy. Uptake was largely independent of socio-demographic and obstetric characteristics. However, the number of antenatal care (ANC) visits was the only factor significantly associated with IPTp-SP uptake. Despite high coverage, qualitative findings revealed important experiential barriers, including perceived side effects, challenges with drug administration, and limited understanding of IPTp-SP may affect adherence and completion of recommended doses.

### Recommendations

At the national level, programme managers should prioritize strategies that promote early initiation and consistent attendance of antenatal care (ANC), given its strong association with IPTp-SP uptake. At the sub-national level, Regional and District Health Directorates should intensify community-based health education and outreach to improve ANC retention and address gaps in service utilization. Additionally, health facility managers and frontline ANC providers should enhance patient-centered counselling by clearly communicating the benefits of IPTp-SP, addressing concerns about side effects, and improving provider–client interactions to foster trust and adherence. These coordinated efforts will be critical to optimizing IPTp-SP uptake and improving maternal and neonatal health outcomes in Ghana and other malaria-endemic settings.

## Author contributions

GA: Conceptualization, Supervision, Methodology, Writing – review & editing, Resources

DTA: Conceptualization, Methodology, Data curation, Validation

NMG: Methodology, Data curation, Writing – review & editing

PM: Methodology, Writing – review & editing, Data curation

IK: Visualization, Data curation, Writing – original draft, Formal analysis, Software, Validation

GYGY: Methodology, Data curation, Writing – review & editing

KJC: Writing – original draft, Project administration, Methodology, Investigation

BB: Investigation, Project administration, Data curation, Writing – review & editing

SC: Writing – original draft, Formal analysis, Software, Visualization, Writing – review & editing

## Acknowledgements

The authors are grateful to the Western Regional Health Directorate and District Health Directorates of Ellembelle, Jomoro, and Prestea-Huni Valley, and the management and staff of the participating health facilities for their support during the study. We sincerely thank all the postpartum women who generously shared their time and experiences. We also acknowledge the contributions of data collectors and field supervisors whose efforts made this study possible.

## List of Abbreviations

ANC: Antenatal care
CWC: Child welfare clinic
DOT: Directly observed therapy
GHS: Ghana Health Service
IPTp: Intermittent preventive treatment in pregnancy
IPTp-SP: Intermittent preventive treatment in pregnancy with sulfadoxine–pyrimethamine
MIP: Malaria in pregnancy
NMEP: National Malaria Elimination Programme
ODK: Open Data Kit
SP: Sulfadoxine–pyrimethamine
SSA: Sub-Saharan Africa
WHO: World Health Organization

## Data availability

All data supporting the findings of this study are available within the paper

